# Single-Arm, Open-Label Phase 2 Trial of Preemptive Methylprednisolone to Avert Progression to Respiratory Failure in High-Risk Patients with COVID-19

**DOI:** 10.1101/2021.03.08.21253117

**Authors:** Fernando Cabanillas, Javier Morales, José G. Conde, Jorge Bertrán-Pasarell, Ricardo Fernández, Yaimara Hernandez-Silva, Idalia Liboy, James Bryan-Díaz, Juan Arraut-Gonzalez

**Affiliations:** University of Puerto Rico School of Medicine; Clinical Research Puerto Rico; University of Puerto Rico Medical Sciences Campus; San Juan Bautista School of Medicine; Auxilio Mutuo Hospital

## Abstract

**Introduction:** Covid-19 is a triphasic disorder first typified by a viral phase that lasts from the first onset of symptoms until seven days later. This is followed by a second and third phase, initially characterized by the appearance of lung infiltrates, followed in 20% by respiratory failure. The second phase is usually heralded by an elevation of serologic inflammatory markers including CRP, ferritin, IL-6, LDH as well as D-dimers. Approximately 20% proceed to the second phase and are usually then treated with dexamethasone, provided they are oxygen-dependent since these are the only cases that benefit from dexamethasone. If we had objective criteria to predict this 20% that develop severe illness, they could preemptively be treated with steroids. In this exploratory study we investigated the early use of preemptive steroids in the setting of early disease, in high-risk non-oxygen dependent cases.

**Methods:** Eligible patients were those 21 years or older with a diagnosis of Covid-19 and oxygen saturation ≥91%. For patients to be classified as high-risk, they had to exhibit two or more of the following abnormalities 7-10 days after first symptom: IL-6 ≥ 10 pg/ml, ferritin > 500 ng/ml, D-dimer > 1 mg/L (1,000 ng/ml), CRP > 10 mg/dL (100 mg/L), LDH above normal range lymphopenia (absolute lymphocyte count <1,000 /µL), oxygen saturation between 91-94%, or CT chest with evidence of ground glass infiltrates. Primary endpoint was progression to respiratory failure. CALL score method was used to predict the expected number of cases of respiratory failure. High risk patients received methylprednisolone (MPS) 80 mg IV daily x 5 days starting no earlier than seven days from first onset of symptoms. The primary endpoint was progression to hypoxemic respiratory failure defined as PaO2 <60 mm Hg or oxygen saturation ≤90%. Secondary endpoints included survival at 28 days from registration, admission to intensive care and live discharge from the hospital. Change in levels of inflammatory markers and length of hospitalization were also assessed.

**Results:** In 76 patients, the expected number with respiratory failure was 30 (39.5%), yet only 4 (5.3%) developed that complication (p=.00001). Survival at 28 days was 98.6%.

Improvement in inflammatory markers correlated with favorable outcome.

**Conclusions:** Our results are encouraging and suggest that this approach is both effective and safe.

## Introduction

Covid-19 is a triphasic disorder first typified by a viral phase that lasts from the first onset of symptoms until approximately 7 days later. This is followed by a second phase considered as the inflammatory stage, characterized first by the appearance of lung infiltrates, which can result in hypoxemia^1-4^. This second phase is usually heralded by an elevation of serologic inflammatory markers such as C-reactive protein (CRP), ferritin, interleukin-6 (IL-6)^5^, LDH as well as D-dimers^4,6,7^. In a smaller subset of cases this is followed by a third phase consisting of hyperinflammation that leads to the cytokine release syndrome or cytokine storm, which causes Acute Respiratory Distress Syndrome (ARDS)^1,2^. Mortality secondary to Covid-19 is usually related to this latter complication.

Approximately 20% of patients will proceed to the second phase. This rate is a crude estimate that can vary according to several prognostic factors^3,4,7^. Currently, there is no reliable and objective method to accurately predict the 80% that are cured spontaneously without any treatment, vis-à-vis those 20% who develop severe illness. There is consensus that those presenting with severe illness characterized by hypoxemia, should be managed in the inpatient setting and treated with dexamethasone ^8^. However, according to the RECOVERY trial, only patients with severe illness requiring oxygen administration in the hospital, benefit from that treatment. Those who are non-oxygen dependent, not only fail to benefit, but could actually be harmed by the use of dexamethasone ^8^.

In this study, we wanted to explore the early use of preemptive steroids in less severe cases that are not oxygen-dependent. However, there are no proven objective criteria to prospectively identify high-risk cases who could be candidates for such treatment.

We hereby report on the results of a Phase II clinical trial whose major aims were to identify non-oxygen-dependent patients with less severe Covid-19, but at high-risk of progressing to hypoxemic respiratory failure, and to evaluate the effect of preemptive treatment with corticosteroids in such cases.

We prospectively divided Covid-19 patients by ranking them into high and low-risk, according to blood-based biomarkers of inflammation, as well as other clinical features. This report is one of two components of a phase II pilot exploratory study designed, in part, to determine if early preemptive therapy with MPS can avert the cytokine storm. The first component regarding the subgroup of low-risk cases is being reported separately ^9^. The current manuscript exclusively focuses on the management of high-risk cases.

## Materials and Methods

### Investigational plan

The study was registered in clinicaltrials.gov as NCT04355247 and approved by the local Institutional Review Board (IRB) at Auxilio Mutuo Hospital in San Juan, Puerto Rico. The informed consent form approved by the local IRB was discussed with each patient and signed prior to registration. The study was conducted at Auxilio Mutuo Hospital in San Juan, Puerto Rico. Patients were recruited between April and December 2020.

The original plan was to enter a total of 100 patients with the expectation that at least 20 would-be high-risk cases eligible for therapy with MPS and 80 would be low-risk, eligible for monitoring without therapy. This expectation was based on the available literature data, which describes a 20% chance for patients with Covid-19 to develop severe disease associated with respiratory failure^3,6^. We later decided to expand this pilot study to include at least 200 patients.

### Eligibility

Eligible patients were those 21 years or older with a diagnosis of Covid-19 established by means of either the PCR molecular test (97% of cases registered) or with the rapid serologic test in the context of typical symptoms and/or ground glass infiltrates in the chest CT (3% of cases). There was no top age limit for entry.

Excluded from entry were those already in acute respiratory failure defined as oxygen saturation ≤90%. Other exclusion criteria included anyone who was chronically oxygen dependent, or who had long standing history of severe COPD. Patients receiving tocilizumab, convalescent plasma, or prednisone 20 mg daily or more, or equivalent, prior to entering the study, were also excluded.

### Definition of high-risk cases

For patients to be classified as high-risk, they had to exhibit two or more of the following abnormalities between days 7-10 after their first symptom: IL-6 ≥ 10 pg/ml, ferritin > 500 ng/ml, D-dimer > 1 mg/L (1,000 ng/ml), CRP > 10 mg/dL (100 mg/L), LDH above normal range, lymphopenia (absolute lymphocyte count <1,000 /µL), oxygen saturation between 91-94%, or CT chest with evidence of ground glass infiltrates. These markers were selected based on published data which have shown a strong association with outcome ^3,4,7^. When the CALL score method to predict prognosis was published ^10^, we amended the protocol to apply this score with the purpose of allowing a more precise estimate of the expected number of cases that would develop respiratory failure. The CALL Score method considers the presence of comorbidities, age, LDH level and lymphopenia to assign a prognostic score^10^. The higher the score, the worse the prognosis. It includes a nomogram which can be used to predict each patient’s risk of progression to respiratory failure.

### Therapy

Patients classified as high-risk were treated with methylprednisolone (MPS) 80 mg IV daily x 5 days starting no earlier than 7 days from first onset of symptoms. Treatment was never started within the first 6 days of illness to avoid prolonging the viral phase by delaying clearance of the virus. Later, the protocol was amended to allow use of a higher dose in morbidly obese patients who were given 160 mg daily x 5 days. This was done in two patients.

Simultaneous treatment with the following drugs was not encouraged but was allowed: hydroxychloroquine, azithromycin, vitamin C, doxycycline, colchicine, zinc and ivermectin. Treatment with tocilizumab was not allowed at time of initiation of MPS but in those cases responding sub-optimally after the third day of MPS, the protocol did not preclude its use. After the 6^th^ patient was registered, we started delivering MPS treatments at the patient’s household by means of a home-health care agency, provided they were stable enough. Of 70 patients eligible for home-based therapy, 58 (83%) received such treatment at home.

Patients were asked to check their oxygen saturation three times per day by means of pulse oximetry and to report any value less than 94% during the first two weeks after entry. They were also instructed to immediately report any unexpected change in their clinical condition. Outpatients were also contacted daily for 28 days to inquire about their condition.

Of the 21 patients who required hospitalization, 16 of these were already hospitalized at the time therapy with MPS was started, because they either had mild hypoxemia or required management of some other Covid complication or co-morbidity already existent at the time they were entered in the study. There were another five admitted following home-based therapy because of respiratory failure in two, dyspnea without respiratory failure in another two, and profound weakness in one. Four of the 21 cases were admitted to the medical intensive care unit (MICU).

### Outcome Measures

Primary endpoint was either progression to hypoxemic respiratory failure defined as an oxygen saturation of ≤ 90% or p02 <60

Secondary endpoints included:

1. Survival at 28 days from registration.
2. Admission to medical intensive care unit (MICU).
3. Live discharge from the hospital.
4. In addition, measurement of inflammatory markers was repeated at day 7 of the study in order to compare with pre-treatment values, and length of hospital admission was also calculated.

Changes in markers were classified into two major categories:

1- normal Pre MPS to normal Post MPS or high pre MPS to normal Post MPS; for ALC the changes were classified as normal Pre MPS to normal Post MPS or low Pre MPS to normal Post MPS.
2- High Pre MPS to high Post MPS or normal pre MPS to high Post MPS; for ALC the changes were classified as low Pre MPS to low Post MPS or normal Pre MPS to low Post MPS.

### Statistical Analysis

Medians and interquartile ranges (IQR’s) were used to describe distributions of continuous variables, and proportions to describe distributions of categorical variables. IQR’s are reported as lower and upper limits of the IQR (i.e., third and first quartiles of distributions). This format not only provides information on the IQR, but also indicate location of the central 50% of distributions relative to the median. The Wilcoxon-Mann-Whitney test was used for testing hypotheses about the difference between distributions of continuous variables, and Fisher’s exact chi-square test for categorical variables. The pairwise sign test was used to assess changes in proinflammatory markers after treatment. The binomial test was used to compare the number of observed cases of respiratory failure to the number expected as predicted by the CALL Score ^11^.

## Results

We initially enrolled 213 patients, 5 of which were not evaluable because consent was withdrawn (n=3), PCR for Covid-19 was negative (n=1) and no blood markers were performed due to suicidal attempt (n=1). Of the 208 evaluable cases, 76 were categorized as high-risk. Median follow-up for the high-risk cases was 3,116 person days (range 1,140-10,488), IQR was 2,622 with 25^th^ percentile =34 and 75^th^ percentile=74.25.

Table 1 depicts the demographics of the whole sample including low as well as high-risk cases. The details of this table have been discussed in another manuscript that deals exclusively with the low-risk cases ^9^.

**Table 1.** Demographics.

| Feature | Low risk (N=132) | % | High-risk (N=76) | % | P |
| --- | --- | --- | --- | --- | --- |
| Gender |  |  |  |  |  |
| Male | 62 | 47 | 40 | 52.6 | 0.36 |
| Female | 70 | 53 | 36 | 47.3 |  |
| Median age (IQR*) | 45<br>(35.5,57) | - | 60<br>(50,72) | - | .00001 |
| Median # comorbidities per patient (IQR) | 1<br>(0,1) | - | 1<br>(1,2) | - | .00001 |
| CALL Score: |  |  |  |  |  |
| 4-6 (Class A) | 64 | 48.5 | 14 | 18.4 | .0001 |
| 7-9 (Class B) | 62 | 47.0 | 36 | 47.3 |  |
| 10-13 (Class C) | 3 | 2.2 | 26 | 34.2 |  |
| Missing information | 3 | 2.2 | 0 | 0 |  |
| O<sup>2</sup> saturation 91-94% | 2 | 1.5 | 23 | 30.2 | .00001 |
| Symptoms: |  |  |  |  |  |
| Fever | 49 | 37.1 | 48 | 63.2 | .0001 |
| Dyspnea | 21 | 15.9 | 29 | 38.2 | .004 |
| Myalgia | 84 | 63.6 | 49 | 64.4 | 1.0 |
| Diarrhea | 55 | 41.7 | 33 | 43.4 | 0.88 |
| Anosmia | 67 | 50.7 | 34 | 44.7 | 0.47 |

### Respiratory Failure

After applying the CALL Score, to the 76 high-risk patients, the expected number of cases of respiratory failure was 30 (39.5%). However, after treatment with MPS, only 4 (5.3%) developed that complication (p=.00001). When CALL score was applied to all cases including low-risk and high-risk features, 206 cases had all the information necessary to calculate the score and the projected respiratory failure rate was 20.9%.

Tocilizumab was administered to two patients who after 5 days of MPS had not improved satisfactorily regarding their symptomatology or oxygen saturation without respiratory failure. It was also given to another two of four patients who had already been counted as respiratory failures. This decision was made in combination with the IL-6 pre-treatment levels which were elevated at 182.8, 70.0, 65.2 and 12.26 pg/ml. All four improved clinically within 24 hours, although one later deteriorated and died. In an additional three cases, treatment with MPS was extended beyond five days due to suboptimal improvement in fever or dyspnea without respiratory failure.

### Difference Between Markers Pre and Post MPS

The change in all markers studied pre and post MPS is shown in table 2. There was a statistically significant difference in CRP, LDH, IL-6, ferritin and absolute lymphocyte count (ALC) before and after treatment. These differences consisted of a drop in markers post-treatment except for ALC which showed an increase (Table 2 and Figure 1). In 17 cases the increase in ALC resulted in complete resolution of lymphopenia.

**Table 2.** Change in Markers (Post-Treatment with Methylprednisolone minus Pre-Treatment)

| **Marker** | **Median difference (IQR)** | **Total patients** | **Increased N (%)** | **Decreased N (%)** | **p-value** |
| --- | --- | --- | --- | --- | --- |
| CRP (mg/dL) | -0.85<br>(-5.20, 0.08) | 73 | 19 (26.0) | 49 (67.1) | < 0.001 |
| LDH units | -44<br>(-100.75, 2.25) | 72 | 19 (26.4) | 53 (73.6) | < 0.001 |
| IL-6 (pg/ml) | -9.00<br>(-15.74, 0.10) | 66 | 15 (22.7) | 49 (74.2) | < 0.001 |
| Ferritin (ng/ml) | -57.3<br>(-174.3, 0.4) | 71 | 18 (25.4) | 52 (73.2) | < 0.001 |
| D-dimer (mg/L) | .02<br>(-0.51, 0.41) | 69 | 30 (43.5) | 36 (52.2) | 0.54 |
| ALC per mL | 542<br>(169, 1,102) | 75 | 62 (82.7) | 13 (17.3) | < 0.001 |

**Figure 1.**
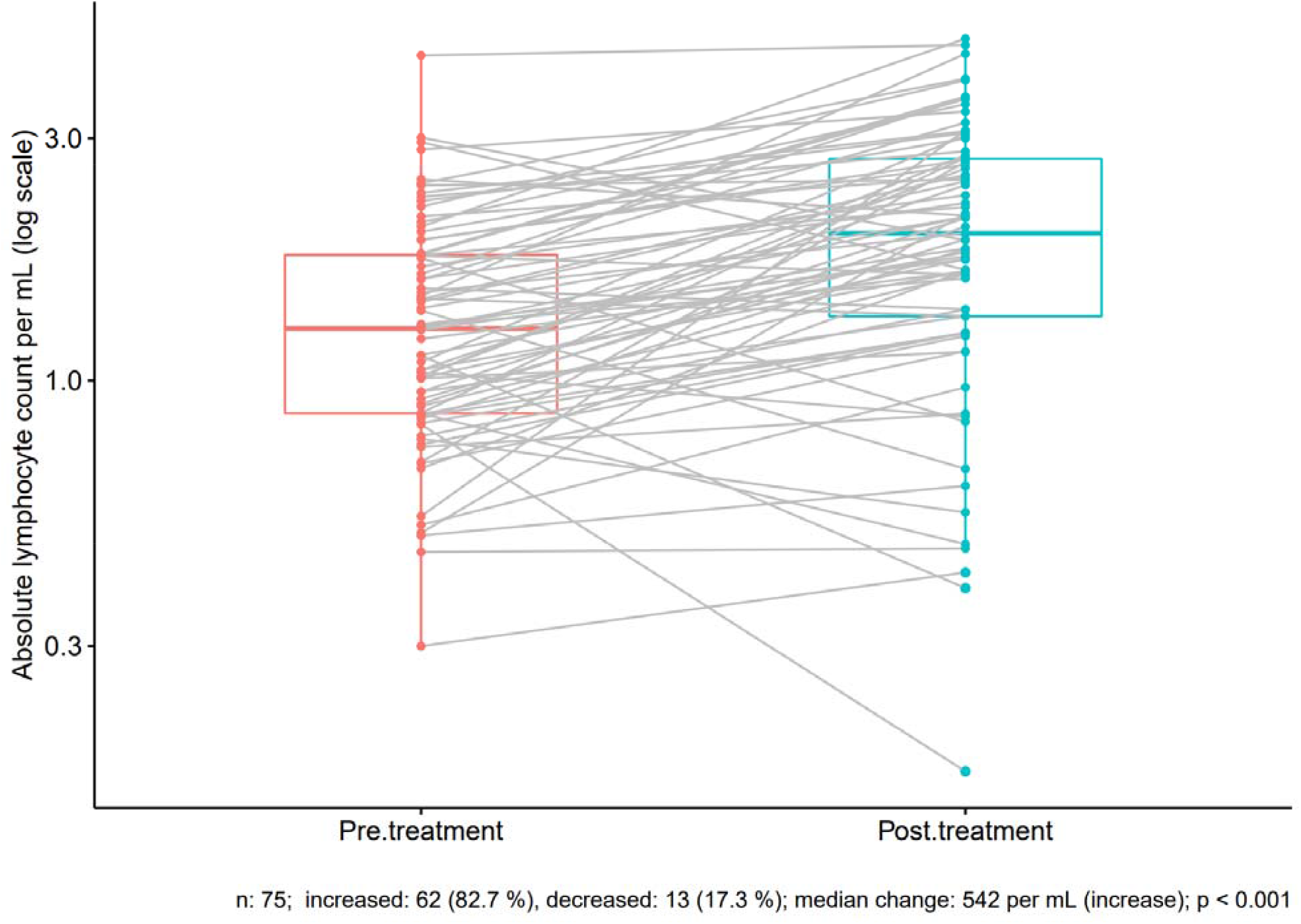
Change in ALC After Treatment with Methylprednisolone.

### Survival, admission to MICU and live discharges

There was one death within 28 days of registration on study, for a 28-day survival rate of 98.6%.. Another patient died on day 57 because of complications related to COVID-19. There were five patients who required admission to MICU at some point during their illness.

Once we started delivering treatment at the patient’s household, which happened eight days after the protocol was initiated, a total of 14 required hospitalization, nine at the time they were first seen and five others who started treatment at home but had to be admitted subsequently. Twelve were discharged alive after a median of 10 days of hospitalization while the other two died in the hospital.

### Changes in blood markers and their association with clinical outcome

We then examined the association between clinical outcome and the change in pre to post-treatment values in pro-inflammatory blood markers as well as with ALC, as shown in table 3

**Table 3.** Association Between Change in Blood Parameters at Day 7 Post MPS Treatment and Clinical Endpoints.

| Change in CRP from pre-treatment to post-treatment | Total died, respiratory failure or MICU admission |  |  | P |
| --- | --- | --- | --- | --- |
|  | Yes | No | Total |  |
| Normal->normal or high->normal | 0 (0) | 45 (100%) | 45 | .00001 |
| High->High or Normal->high | 10 (35.7%) | 18 (64.3%) | 28 |  |
| Change in LDH from pre-treatment to post-treatment | Yes | No | Total | P |
| Normal->normal or high->normal | 2 (3.9%) | 49 (96%) | 51 | .0005 |
| High->High or Normal->high | 8 (38.1%) | 13 (61.9%) | 21 |  |
| Change in Ferritin from pre-treatment to post-treatment | Yes | No | Total | P |
| Normal->normal or high->normal | 1 (3.6%) | 27 (196.4%) | 28 | .14 |
| High->High or Normal->high | 7(16.3%) | 36 (83.7%) | 43 |  |
| Change in D-dimers from pre-treatment to post-treatment | Yes | No | Total | P |
| Normal->normal or high->normal | 0 (0) | 28 (100%) | 28 | .004 |
| High->High or Normal->high | 10(24.4%) | 31 (75.6%) | 41 |  |
| Change in IL-6 from pre-treatment to post-treatment | Yes | No | Total | P |
| Normal->normal or high->normal | 2 (3.8%) | 51 (96.2%) | 53 | .009 |
| High->High or Normal->high | 4 (33.3%) | 8 (66.7%) | 12 |  |
| Change in ALC* from pre-treatment to post-treatment | Yes | No | Total | P |
| Normal->normal or low->normal | 3 (4.7%) | 60 (95.2%) | 63 | <.00001 |
| Low->Low or Normal->Low | 8 (67%) | 4 (33.3%) | 12 |  |

\*ALC= Absolute Lymphocyte Count

Table 3 portrays the various patterns from pre to post MPS levels and their association with clinical outcome, defined as number of unfavorable events or endpoints which included death, admissions to MICU and respiratory failure. There was a statistically significant association between CRP, LDH, D-dimers and IL-6 (Table 3) and the combined unfavorable endpoints. However, the most noticeable association observed was with the post treatment changes in ALC. As expected, when the pattern was from normal to normal or low to normal, only 4.7% of 63 cases had an unfavorable outcome while there were 66.6% of 12 cases who met at least one of the unfavorable endpoints if the pattern was from low to low or normal to low ALC (p<.0001).

A pre-treatment ALC ≤1,000 has been previously associated with a high rate of respiratory failure ^10^, so we attempted to confirm this finding. Table 4 examines the association of pre-treatment ALC with respiratory failure and shows that in contrast with the difference between pre and post treatment ALC, there is no statistically significant association at any of the levels examined for baseline ALC.

**Table 4.** Association Between Various Pre-treatment ALC Levels and Clinical Outcome.

| **ALC /μL at Baseline** | **N=76** | **Respiratory Failure** | **P value** |
| --- | --- | --- | --- |
| $\geq 500$ | 73 | 3 (3.9%) | .15 |
| < 500 | 3 | 1 (33.3%) |  |
| $\geq 750$ | 64 | 3 (4.6%) | .5 |
| < 750 | 12 | 1 (8.3%) |  |
| $\geq 1,000$ | 50 | 2 (4%) | 0.6 |
| < 1,000 | 26 | 2 (7.7%) |  |

ALC= absolute lymphocyte count
Pre-treatment refers to day 7-10 post first symptom.
day 7 CBC was not done in 1 patient.

### Toxicity

The only serious adverse reaction observed was in two diabetic patients (one type II and the other one type I), whose blood sugar while on MPS increased to the point that an addition or increase of insulin was necessary. Otherwise, MPS was well tolerated.

## Discussion

The use of corticosteroids in the management of Covid-19 has been criticized because of the legitimate concern regarding the use of immunosuppressive medications that may cause a delayed viral clearance and increase the risk of secondary infections. However, the data available against the use of steroids ^12^ was derived in great extent from trials in which they were used either too early during the viral phase, thus placing patients at risk of worsening their infection, or too late, at which time their efficacy was compromised.

After recognizing the role of excessive inflammation as an important factor in the pathogenesis of respiratory failure in Covid-19, dexamethasone was successfully introduced and tested in the RECOVERY trial for hospitalized patients ^8^. Later, high dose MPS ^13^ was used in the Netherlands by Ramiro et al in a clinical trial designed exclusively for the management of an established cytokine storm. As shown in these two trials, corticosteroids are effective. However, in cases with advanced presentations, the mortality can be reduced but is still considerable ^8,13^.

In early 2020, Dr. Angel Atienza, from Hospital Doctor Peset, Valencia, Spain proposed the use of a brief course of MPS early during the illness, right at the end of the viral phase, and shortly before the second or inflammatory phase of the disease (personal communication). The aim was to utilize it as preventive therapy for patients at high risk of entering the second phase. He proposed to apply it to non-oxygen dependent cases with high-risk clinical features identified by certain defined abnormalities in their blood parameters. In the RECOVERY trial these oxygen-independent patients failed to benefit, or might have even been harmed by the 10 day course of dexamethasone ^8^.

According to the published medical literature ^4,14^, the number of cases with Covid-19 expected to develop respiratory failure is roughly 20%, which is equivalent to 42 of the 208 patients entered on our study. This closely matches the projection of 43 expected cases estimated by applying the CALL score. In the 76 high-risk cases we treated, a respiratory failure rate of 39.5% was expected according to their CALL score, but the observed rate was much lower, 5.3% (p<.00001). None of the 132 low-risk cases developed respiratory failure. We conclude that this approach has the potential of averting the cytokine storm commonly seen in this disorder, if applied to high-risk cases after day 7-10, but never earlier than 7 days after the first symptom.

In the RECOVERY trial, oxygen-independent patients fared poorly when treated with dexamethasone, yet our findings in oxygen-independent cases treated preemptively with MPS are encouraging. These data strongly suggest that this novel preemptive approach is not only effective but also safe.

Particularly interesting was the association we observed between clinical outcome and changes in pre-treatment CRP, LDH, D-dimers, IL-6 compared with the day 7 post MPS values (table 3), which suggests that MPS might have reduced inflammation, thus decreasing complications including respiratory failure.

The weak association we observed between pre-treatment ALC and development of respiratory failure (table 4), although unexpected, perhaps should not be surprising. It is a well-known fact that when newer and more effective treatments are introduced, these can alter or totally abolish well-established prognostic factors ^15^.

The increase in ALC observed in 63 cases is consonant with our interpretation that this treatment could have brought about this improvement which correlates well with a lower rate of respiratory failure, while those whose day 7 ALC post-MPS failed to improve, or dropped below normal, tended to fare less well. A similar pattern was observed in day 7 post MPS levels of CRP, LDH, D-dimers, and IL-6 (table 3), again suggesting a beneficial effect of this treatment.

Possible confounding factors include the use of tocilizumab in two patients and the additional doses of MPS in another three cases who were not responding as expected. However, even if we do not consider these five cases as successfully treated, the results still would favor MPS since a total of 9 (11.8%) would be considered as failures compared with the 30 expected cases (39.5%), p=.0002.

Although the data generated by this exploratory trial appear strong and compelling, confirmation of these findings in an independent study would be highly desirable.

## Data Availability

Available on request

The authors report no conflicts of interest.

## Acknowledgements

This study was supported in part by:

1. Puerto Rico Coalition for Clinical Investigation which provided support for data management.
2. This project was partially funded by RCMI Grant U54 MD007600, NIMHD, NIH. The content of this report is solely the responsibility of the authors and does not necessarily represent the official views of the National Institutes of Health.
3. Best Option Company provided treatment with methylprednisolone at patients’ households free of cost.

